# The Paisa Kids Study: Rationale, Methods, and Initial Characterization of a Youth Cohort at Transdiagnostic High Risk for Serious Mental Illness

**DOI:** 10.64898/2026.10.05.26364818

**Authors:** Jennifer K. Forsyth, Johanna Valencia-Echeverry, Carrie E. Bearden, Nelson B. Freimer, Loes M. Olde Loohuis, Jason J. Karalis, Victor I. Reus, Javier I. Escobar, Carlos A. Lopez-Jaramillo, Juan David Palacio-Ortiz

**Affiliations:** Department of Psychology, University of Washington, Seattle, WA; Institute for Public Health Genetics, University of Washington, Seattle, WA; Research Group in Psychiatry (GIPSI), Department of Psychiatry, Faculty of Medicine, University of Antioquia, Medellín, Colombia; Center for Neurobehavioral Genetics, Semel Institute for Neuroscience and Human Behavior, David Geffen School of Medicine, University of California Los Angeles, Los Angeles, CA; Department of Human Genetics, University of California Los Angeles, Los Angeles, CA; Department of Psychiatry and Behavioral Sciences, University of California San Francisco, San Francisco, CA; Department of Psychiatry, Rutgers University, New Brunswick, NJ

**Author notes:** **Corresponding Author:** Jennifer Forsyth, PhD, Department of Psychology, University of Washington, Institution, 3751 W Stevens Way NE, Seattle, Washington, United States, 98195.

## Abstract

**Objective:** The Genetic Risk for Serious Mental Illness and Development (“Paisa Kids”) Study is establishing a longitudinal cohort of 3,000 youth enriched transdiagnostically for risk for serious mental illnesses (SMI; i.e., schizophrenia, bipolar disorder, and severe depression) from the Paisa region of Colombia. Its objective is to clarify the early clinical, neurobehavioral, genetic, and environmental profiles associated with later SMI and distinguish transdiagnostic from disorder-specific pathways. This paper describes the study infrastructure, recruitment streams, and baseline assessment battery, and characterizes the first two-thirds of the cohort.

**Method:** Youth aged 6–15 years are being recruited. Youth recruited for high SMI risk either have a first or second degree adult relative with a SMI diagnosis or a psychiatric diagnosis themselves identified through electronic medical records (EMRs) associated with later SMI. Lower-risk youth have a relative without a SMI diagnosis. Baseline measures include psychiatric and developmental history, neurobehavioral functioning, environmental exposures, and genome-wide genotyping. Psychiatric EMRs provide longitudinal follow-up.

**Results:** Among 2,384 participants enrolled thus far (mean age=11.2 years, 45.0% female), 2,211 were recruited for high SMI-risk and 173 for lower SMI-risk. Overall, 73.2% met criteria for a history of one or more psychiatric disorders. Logistic regression comparing rates of diagnoses and ACEs between recruitment streams showed elevated rates of depressive and anxiety disorders, attention-deficit/hyperactivity disorder, oppositional defiant disorder, and several ACEs among high versus lower SMI-risk youth.

**Conclusion:** Integrating multidomain phenotyping, genomic data, and longitudinal EMR follow-up, the Paisa Kids cohort provides a unique resource for investigating pathways of risk and resilience to SMI.

## Introduction

### Background Rationale

Improving our ability to predict and prevent the onset of serious mental illnesses (SMI), such as schizophrenia (SCZ), bipolar disorder (BP), and severe depression (sev-DEP), remains a critical goal in psychiatry. These disorders are often diagnosed in advanced stages, when suffering has reached significant levels and restoring functioning to premorbid levels is a challenge. Within a clinical staging framework, the onset of major psychiatric symptoms, such as psychosis in SCZ or mania in BP, can be conceptualized as late stages of a progressive trajectory of altered brain development.^1^ In line with this, epidemiological studies indicate that major “adult” psychiatric illnesses evolve in clarity and severity of symptoms over time.^1^ Furthermore, meta-analytic studies suggest that early medication- and/or psychosocial-based interventions can alter disease trajectory in individuals with subthreshold psychotic symptoms, including preventing SMI onset and improving long-term outcomes for at least some individuals.^2,3^ However, limitations of our current psychiatric nosology, including the imprecise boundaries between disorders and undifferentiated and frequently overlapping features of early clinical presentations preceding SMI^4^ remain obstacles to optimizing prediction of distinct SMI and developing targeted interventions to maximize prevention.

To address these obstacles, the “Genetic Risk for Serious Mental Illness and Development” or “Paisa Kids” project was initiated in 2021, with the aim of establishing a cohort of 3,000 youth enriched transdiagnostically for SMI risk for longitudinal follow-up. This study builds upon research infrastructure from large-scale genetic studies of adult SMI in the Paisa region of Colombia, initiated 20 years ago due to the genetic isolate nature of the Paisa population, which can enhance power for genetic discovery.^5^ The majority of the youth cohort is being recruited for high SMI risk because they either: 1) have an adult relative with a SMI diagnosis enrolled in prior studies in the region; or 2) already have a diagnosis as a child in the electronic medical records (EMR) of local psychiatric hospitals that is associated with elevated risk for future SMI onset. An additional subset of youth is being recruited for lower SMI risk because they have an adult relative who enrolled in prior studies and did not have a SMI diagnosis. By obtaining genome-wide genotyping, conducting a comprehensive clinical and neurobehavioral assessment at study entry, and using EMR to derive longitudinal clinical outcomes, initial funding was procured to establish the youth cohort towards the long-term goals of clarifying the clinical, neurobehavioral, genetic, and environmental profiles that can optimize prediction of later SMI onset.

### Infrastructure Supporting the Study

State-of-the-art research and health-service related infrastructure, combined with unique characteristics of the population and sociopolitical history of Colombia make the Paisa population an important population for understanding risk and protective factors for SMI. Thus, the Paisa population of ~9 million people predominates in a densely populated region of the Andean Mountains that is located mainly in Antioquia, Colombia. The population was founded in the 16^th^-17^th^ centuries with the admixture of indigenous women and Spanish colonist men, and includes a small African ancestry contribution from enslaved individuals who were forcibly brought to the region by European colonizers.^6,7^ Owing to geographical isolation imposed by the mountainous terrain, the population had low mobility while undergoing rapid expansion over the last 300 years. The reduced genetic heterogeneity characteristic of such genetic isolates and large genealogical size of many Paisa families^5^ prompted the initiation of genetic studies of SMI in the region over 20 years ago and established critical research infrastructure for the current study^8,9^. In addition, the sociopolitical history of Colombia through the 20th and 21st centuries resulted in elevated exposure for many youth to traumatic and/or adverse experiences relevant for the development of SMI. This includes political violence, the rise of drug cartels in the 1970s to 1990s, and an armed conflict between 1964 and 2016 that resulted in elevated exposure to violence and one of the highest rates of forced internal displacement, globally, for many years.^8,10^ Finally, while socioeconomic levels in Antioquia vary widely and most people in Colombia are in the low and middle-low socioeconomic levels, Colombia is a middle-upper income country overall, and health infrastructure in Antioquia includes EMR that has been available for over 15 years in all health facilities. This facilitates access to clinical data and passive longitudinal monitoring of patients with and without mental disorders, facilitating investigation into risk and protective factors in the development of SMI.

Recruitment and longitudinal EMR-based follow-up of the current “Paisa Kids” cohort is directly enabled by two prior NIH-funded studies of adults with SMI in the region: (1) the “Genetics of Severe Mental Illness” project, also often termed the “Paisa Project,” and (2) the “Latin American Biobank for Large-Scale Genetics Research on Severe Mental Illness” (LAB-SMI) project, also often termed, “Mision Origen.” The “Genetics of Severe Mental Illness*”* project was designed to generate deep-phenotyping (i.e., clinical interviews and assessments of dimensional traits) on 8,000 Paisa adults with SMI, including SCZ spectrum, BP spectrum, and severe-DEP patients, along with 2,000 Paisa adults without SMI, for genetic investigation.^11^ Adults with SMI were recruited from the Clinica San Juan de Dios, Manizales (CSJDM) psychiatric hospital in Manizales, Caldas, which is the main psychiatric hospital for the department (i.e., state) of Caldas, which has a population of ~1 million, and the Hospital Mental Antioquia (HOMO) in Medellin, Antioquia. CSJDM is a private psychiatric hospital while HOMO is one of the largest public psychiatric hospitals in Colombia. EMR at each of these hospitals were established as research databases. The “LAB-SMI” study expanded use of the EMR in Colombia to broader psychiatric hospitals that together treat the majority of SMI patients in the region. LAB-SMI aimed to: 1) create a Latin American DNA biobank of 100,000 Paisa adults, involving 50,000 SMI individuals and 50,000 controls; 2) map associations of common and rare variants with SMI diagnoses, symptoms, and behaviors in the Paisa population; and 3) explore the architecture of EMR-derived phenotypes. This EMR-based phenotyping study established invaluable infrastructure for deriving long-term clinical outcomes for youth enrolled in the current study.

### Current Study Aims

The primary analytic aims of the current “Paisa Kids” study are to:

1. Characterize relationships between clinical syndromes and neurobehavioral traits in childhood.
2. Map relationships between common variant-based genetic risk for SMI, rare damaging variant burden, and psychiatric diagnoses in childhood.
3. Map relationships between common variant-based genetic risk for SMI, rare damaging variant burden, and cognitive, motor, sensory, and psychological markers of functioning.

Additionally, using psychiatric EMR databases to obtain longitudinal outcomes, we aim to explore genetic, clinical, environmental, and neurobehavioral characteristics associated with outcome within 2 years.

These aims were planned to help clarify trans-diagnostic vs. disorder-specific risk profiles in childhood, identify clinical syndromes and neurobehavioral traits in childhood associated with genetic risk for SMI, and quantify the power of genetic versus clinical and neurobehavioral characteristics for predicting proximal psychiatric outcomes. However, clinical outcomes will continue to be extracted from the EMR beyond the initial funding period. Data from this cohort will also be made available to the scientific community through the NIH Data Archive (NDA). Thus, there are many additional questions that can be explored by the broader scientific community in this cohort.

To facilitate dissemination of this data, the current paper therefore describes the recruitment streams and measures collected at baseline assessment, and additionally provides an initial characterization of the first two-thirds of the cohort. This includes a brief comparison of lifetime history of psychiatric diagnoses and exposures to adverse childhood experiences (ACEs) between youth recruited for high versus lower SMI-risk.

## Methods

### Recruitment Streams

The majority of the “Paisa Kids” cohort is being recruited to be at high SMI-risk based on either family history of SMI or because the youth participant themselves has a diagnosis in the EMR of local hospitals associated with increased risk of future SMI (target *n* = 2,700). A subset of the youth are being ascertained for lower SMI risk because they have a relative with no SMI diagnosis in our prior adult-based study (target *n* = 300). These recruitment streams are summarized below.

#### Family-Based SMI High-Risk Stream

One thousand, five hundred children are being ascertained for elevated SMI risk because they have a first- or second-degree relative with a SCZ spectrum, BP spectrum, or severe-DEP diagnosis (target *n* = 500 each) in the adult “Genetics of Severe Mental Illness” study.

#### EMR-Based High SMI-Risk Stream

One thousand, two hundred children are being ascertained for elevated SMI risk because they have a diagnosis in the EMR of HOMO or CSJDM that was associated with a later SMI diagnosis, based on analyses of the CSJDM EMR. In particular, youth with the following diagnoses in the EMR are being recruited: attention deficit hyperactivity disorder (ADHD), conduct disorder, mild-moderate intellectual disability (ID), autism spectrum disorder (ASD), generalized anxiety disorder (GAD), severe stress reaction, affective disorder, not otherwise specified, or major depressive disorder (MDD; target *n* = 150 per diagnosis category).

#### Family-Based Low SMI-Risk Stream

300 youth are being ascertained for lower SMI risk because they have a first or second-degree adult relative who did not have a SMI diagnosis in the “Genetics of Severe Mental Illness” study.

Exclusion criteria across recruitment streams were kept minimal to: (1) significant intellectual impairment (IQ estimate < 55); (2) known chromosomal aneuploidy such as Trisomy 21; and (3) history of significant brain trauma or disorder (e.g., brain tumor, head injury resulting in coma). These criteria were used to exclude children who were at high likelihood of being unable to complete informed assent or provide valid assessment data.

This ascertainment strategy was designed to enrich the cohort for elevated SMI risk while ensuring variation in the extent of genetic risk to optimize statistical power. For the family-based recruitment streams, study participation was limited to four youth family members per adult with a SMI diagnosis or without a SMI diagnosis in the prior “Genetics of Severe Mental Illness” study. Informed consent was obtained from the parent or primary caregiver and assent was obtained from the minor.

### Baseline Assessment Measures

In the following, we describe the assessment battery administered at baseline and the demographic and clinical characteristics of the initial approximate two thirds of the sample (*n* = 2,384). The battery is summarized in Table 1, and briefly described below, with additional details provided in the supplement. The battery was selected to balance a comprehensive survey of clinical, developmental, environmental, and neurobehavioral domains relevant to the development of SMI, while minimizing participant and assessor burden. Specific measures selected were chosen to parallel other large-scale general population and high SMI-risk cohorts when possible, and when Spanish versions were available (e.g., the North American Longitudinal Study (NAPLS)^12,13^ and United States-based Adolescent Brain and Cognitive Development Study (ABCD)^14^).

**Table 1.** Summary of Assessment Battery.

| Broad Domain | Sub Domain | Instrument |
| --- | --- | --- |
| Cognitive | **IQ Estimate** | • WISC-IV - C |
|  | **Executive Function** | • NIH-T Dimensional Change Card Sort Test - C |
|  | **Attention** | • NIH-T Flanker Inhibitory Control and Attention Test - C |
|  | **Episodic Memory** | • NIH-T Picture Sequence Memory Test - C |
|  | **Language** | • NIH-T Picture Vocabulary Test - C |
|  | **Processing Speed** | • NIH-T Pattern Comparison Test - C |
|  | **Reading** | • NIH-T Oral Reading Recognition Test - C |
|  | **Working Memory** | • NIH-T Word List Sorting Test - C |
|  | **Verbal Learning** | • Rey Auditory Verbal Learning (3 Immediate Memory Trials Administered) - C |
| Sensory | **Auditory Function** | • NIH-T Words-in-Noise Test - C |
| Motor | **Dexterity** | • NIH-T 9-Hole Pegboard Dexterity Test - C |
| Psychological Well-Being | **Clinical Diagnoses + Social, Activities, School, Overall Functioning** | • K-SADS-COMP Diagnostic Clinical Interview - Adol/P<br>• Global Assessment of Functioning (Clinician Rating) |
|  | **Dimensional Psychopathology** | • Child Behavior Checklist - P |
|  | **Personality** | • Big Five Questionnaire - Short Form - C |
|  | **Anhedonia** | • KSADS Item from Depression Module - Adol/P<br>• Adolescent Anhedonia Scale - Adol |
| Other Environment/ Medical History | **Environmental** | • Adverse Childhood Experiences Questionnaire - Adol/P<br>• KSADS Trauma Screener Items - Adol/P<br>• Questionnaire of Unpredictability in Childhood - Adol/P<br>• Child Promotive Factors - Adol/P |
|  | **Developmental History** | • Developmental Milestones - P<br>• Lewis-Murry Obstetric Complications Scale - P |
|  | **Medical & Psychiatric History** | • History of Head Injury - P<br>• Psychiatric Service History (from KSADS) - Adol/P<br>• Current Psychiatric Medication (from KSADS) - Adol/P<br>• Family Psychiatric History (from KSADS) - Adol/P |

\*Abbreviations: NIH-T = NIH Toolbox; C = All Children; Adol = Adolescents Only; P = Parent

The study complied with regulations for human subjects research, as mandated by Colombian laws and the Declaration of Helsinki, and was approved by the research ethics committees at the University of Washington and HOMO hospital.

#### Demographics

A semi-structured interview was conducted with the parent or primary caregiver to obtain demographic and socioeconomic-related information for the youth participant and their family. Socioeconomic classification in Colombia is defined based on a system established by Law 142 of 1994 that assigns different rates for public utility services across socioeconomic strata and divides homes into six strata (i.e., 1 to 6). Households in the highest strata (5 and 6) make additional payments that subsidize the services of the lowest strata (1, 2, and 3), while stratum 4 neither receives subsidies nor makes additional contributions. The classification of a home is determined by its physical characteristics and those of its environment, such as the quality of materials, infrastructure, and the type of neighborhood, rather than the personal income of the residents.

#### Mental Health, Behavioral Traits, and Psychosocial Functioning

##### Kiddie Schedule for Affective Disorders and Schizophrenia - Computerized (KSADS-COMP)

The Spanish version of the computer-assisted KSADS-COMP structured clinical interview, which is the most widely used and well-validated diagnostic interview for children and adolescents^15,16^, was used to obtain psychiatric diagnosis history and history of psychiatric services at study entry. Primary caregivers and the minor were interviewed separately. A consensus diagnosis integrating both reports was determined by trained study staff (i.e., psychologists and social workers).

The KSADS-COMP was also used to collect information on family psychiatric history (i.e., first- or second-degree relative history of depression, psychotic symptoms, manic episodes, alcohol and drug addiction problems, or attempted suicide).

#### Child Behavior Checklist (CBCL)

Dimensional ratings of emotional and behavioral function in the past 6 months were obtained via the CBCL, which is a widely used and well-validated 138-item caregiver-report measure for children and adolescents aged 6 to 18.^17,18^ Adolescents, aged 12 and up, additionally completed the self-report version of the scale.

##### Anhedonia Scale

Anhedonia was measured using the 14-item self-report anhedonia scale for adolescents which quantifies social versus physical anhedonia severity in the past two weeks.^19^

##### Big Five Questionnaire for Children (BFC-SF)

Growing evidence suggests that personality traits may be important predictors of later psychopathology.^20–22^ Personality was therefore assessed with the BFC-SF^23^, which is a 20-item self-report scale that assesses the five dimensions of the Big Five model of personality: openness to experience, conscientiousness, extraversion, agreeableness, and neuroticism.

#### Neurobehavioral Functioning

##### NIH Toolbox for Neurological and Behavioral Functioning (NIH-TB)

Cognitive, sensory, and motor function were assessed via the Spanish version of the NIH-TB^24,25^, which is an expert-curated compilation of assessment measures developed as part of the NIH Blueprint for Neuroscience Research to create a streamlined, “common currency” for assessing neurobehavioral constructs across developmental stages.

Specifically, all subtests from the main NIH-TB cognitive battery were administered, allowing scores for individual cognitive subtests and a total composite score to be generated. The Words-in-Noise Test, which assesses ability to perceive words in a noisy environment, was used to assess sensory function. The 9-Hole Pegboard Test, in which participants sequentially place cylindrical pegs into a 3 by 3 matrix of holes, followed by removing them, was used to assess fine motor ability. See Supplementary Information for details.

##### Rey Auditory Verbal Learning Test

In addition to the NIH-TB cognitive battery, a Spanish version of the Rey Auditory Verbal Learning Test was administered to provide a measure of short-term verbal learning and memory^26^.

##### Wechsler Intelligence Scale for Children - Fourth Edition

The Wechsler Intelligence Scale for Children (WISC) is one of the most widely used measures of intelligence for children. The short-form of the WISC-IV, Spanish edition, involving the Vocabulary, Similarities, Coding, and Matrix Reasoning subscales, was administered to obtain an IQ estimate to screen for significant intellectual impairment (i.e., IQ < 55; exclusion criteria) and facilitate comparison with other studies.^27,28^

#### Environment

##### Adverse Childhood Experiences (ACEs) Questionnaire

ACEs are well-established risk factors for a range of psychiatric disorders.^29^ ACEs were assessed using the 17-item ACE questionnaire, which covers multiple domains including physical, emotional, and sexual abuse, physical and emotional neglect, substance abuse or mental illness in a family member, parental separation or divorce, and domestic violence.^30^

##### Questionnaire of Unpredictability in Childhood

In addition to traditional ACEs, growing evidence indicates that unpredictability in early childhood is associated with increased risk for mental health difficulties.^31,32^ Unpredictability was measured with the 5-item Questionnaire of Unpredictability in Childhood - Spanish - Brief (QUIC-SP-5).^33^

##### Child Promotive Factors

Questions related to promotive factors for youth health across individual-, family-, and neighborhood-levels were adapted from the United States National Survey of Children’s Health Questionnaires for ACEs and Promotive Factors, similar to others.^34^

#### Obstetric, Developmental, and Medical History

##### Lewis-Murray Obstetric Complications Scale

Obstetric complications are established risk factors for SMI, particularly schizophrenia.^35–38^ Thus, obstetric history was assessed using the Lewis-Murray scale, which is a commonly used, 15-item retrospective recall measure of complications related to pregnancy, abnormal fetal growth, and delivery.

##### Developmental and Medical History

Delays in developmental milestones are associated with a number of neurodevelopmental disorders as well as SMI outcomes, particularly schizophrenia, but also potentially bipolar disorder.^39,40^ Caregivers were asked to report what age their child smiled at parents, sat without support, crawled, walked without support, spoke their first word, and spoke in 2-3 word phrases.

#### Genetic Data

DNA was collected via saliva to facilitate participation. Genome-wide genotyping is being completed using Illumina’s Global Screening Array (GSA) II, which covers approximately 660K single nucleotide polymorphism (SNP) markers, optimized for cross-ethnic imputation. Quality control and imputation of genotype data will be performed using the Ricopili pipeline^41^, and initial imputation will be conducted using the TOPMed reference panel.^42^ DNA biospecimens remaining after genotyping will be stored at the NIMH Repository and Genomics Resource (NRGR) for future research.

#### Longitudinal Follow-up

EMR from the psychiatric hospitals, CSJDM in Manizales, and HOMO in Medellin, which are both recruitment sites for the study, will be used to derive longitudinal outcomes for the cohort. Key longitudinal outcomes include diagnoses associated with inpatient or outpatient clinical visits and services associated with worsening symptoms, such as psychiatric hospitalization.

#### Statistical Analyses

Analyses in the current paper were run using R Studio. Descriptive statistics are reported for demographic characteristics, psychiatric diagnostic history, and ACEs for all currently enrolled participants. In addition, logistic regression models, with age and sex as covariates, were used to compare rates of lifetime history of psychiatric disorders (i.e., consensus diagnosis based on caregiver- and child-report on the KSADS-COMP) and ACEs (i.e., derived from the ACE-Q) between youth recruited to be at transdiagnostic high SMI-risk versus lower SMI-risk. P-values for psychiatric disorders and ACEs were corrected for multiple testing, separately, via the Benjamini-Hochberg false-discovery rate (FDR) correction.

### Initial Characterization of Cohort

#### Demographic Characteristics

Table 2 shows the demographic characteristics of the first 2,384 participants (45.0% female) and their caregivers, 2,211 of whom were recruited for high SMI-risk (i.e., 81.9% of high SMI-risk target *n*), and 173 of whom were recruited for lower SMI-risk (i.e., 57.7% of lower SMI-risk target *n*). The average age of the youth at enrollment is 11.2 years (SD = 2.77). Most are at the lowest socioeconomic strata in Colombia, levels 1 and 2 (72.6%). Over half live in a single-parent family. Within the SMI-relative recruitment stream, the largest portion of youth thus far were recruited based on having a family member with a BP diagnosis.

**Table 2.** Sociodemographic characteristics for overall currently enrolled cohort and by serious mental illness (SMI) risk group.

| Variable | Total Sample | High SMI-Risk | Low SMI-Risk |
| --- | --- | --- | --- |
| N (% Female) | 2384 (45%) | 2211 (44.3%) | 173 (53.4%) |
| Mean Age (S.D) | 11.2 (2.77) | 11.2 (2.77) | 10.5 (2.69) |
| Education Level [n (%)] |  |  |  |
| Preschool | 20 (0.8%) | 14 (0.6%) | 6 (3.5%) |
| Primary education (grades 1-5) | 1081 (45.3%) | 981 (44.4%) | 100 (57.8%) |
| Secondary education (grades 6-11) | 1196 (50.2%) | 1132 (51.2%) | 64 (37%) |
| Special education | 8 (0.3%) | 8 (0.4%) | 0 (0%) |
| Out-of-School | 67 (2.8%) | 64 (2.9%) | 3 (1.7%) |
| Unknown | 7 (0.3%) | 7 (0.3%) | 0 (0%) |
| Socioeconomic stratum [n (%)] |  |  |  |
| Lowest level 1 | 592 (24.8%) | 562 (25.4%) | 30 (17.3%) |
| Low level 2 | 1138 (47.7%) | 1061 (48%) | 77 (44.5%) |
| Middle low level 3 | 557 (23.4%) | 500 (22.6%) | 57 (32.9%) |
| Middle high level 4 | 79 (3.3%) | 71 (3.2%) | 8 (4.6%) |
| High levels 5 and 6 | 15 (0.6%) | 14 (0.6%) | 1 (0.6%) |
| Source of family income [n (%)] |  |  |  |
| Family/Spouse/Partner | 627 (26.3%) | 585 (26.5%) | 42 (24.3%) |
| Employed | 858 (36%) | 783 (35.4%) | 75 (43.4%) |
| Self Employed | 812 (34.1%) | 758 (34.3%) | 54 (31.2%) |
| Pension/Insurance | 60 (2.5%) | 58 (2.6%) | 2 (1.2%) |
| Street Vendor | 22 (0.9%) | 22 (1%) | 0 (0.0%) |
| Type of household [n (%)] |  |  |  |
| Single Parent | 1353 (56.8%) | 1278 (57.8%) | 75 (43.4%) |
| Two Parents | 742 (31.1%) | 664 (30%) | 78 (45.1%) |
| Group Home | 123 (5.2%) | 107 (4.8%) | 16 (9.2%) |
| Foster Home | 7 (0.3%) | 6 (0.3%) | 1 (0.6%) |
| In a Shelter | 2 (0.1%) | 1 (0%) | 1 (0.6%) |
| Other | 154 (6.5%) | 152 (6.9%) | 2 (1.2%) |
| Mother's marital status [n (%)] |  |  |  |
| Single | 804 (33.7%) | 754 (34.1%) | 50 (28.9%) |
| Married | 1299 (54.5%) | 1193 (54%) | 106 (61.3%) |
| Separated | 205 (8.6%) | 191 (8.6%) | 14 (8.1%) |
| Divorced | 24 (1%) | 23 (1%) | 1 (0.6%) |
| Widowed | 43 (1.8%) | 41 (1.9%) | 2 (1.2%) |
| Cohabiting | 5 (0.2%) | 5 (0.2%) | 0 (0.0%) |
| Recruitment group [n (%)] |  |  |  |
| Adult Relative With SMI | 1036 (43.5%) | 1036 (46.9%) | 0 (0.0%) |
| Adult Relative Without SMI | 173 (7.3%) | 0 (0.0%) | 173 (100%) |
| Electronic Medical Record | 1175 (49.3%) | 1175 (53.1%) | 0 (0.0%) |
| Relative with SMI [n (%)] |  |  |  |
| Bipolar Disorder | 471 (19.8%) | 471 (21.3%) | 0 (0.0%) |
| Schizophrenia | 213 (8.9%) | 213 (9.6%) | 0 (0.0%) |
| Depression | 339 (14.2%) | 339 (15.3%) | 0 (0.0%) |

#### Psychiatric Disorders History

Table 3 shows the lifetime history rates for psychiatric diagnoses overall across the currently enrolled cohort, as well as split by youth recruited for high-versus lower-SMI risk. Overall, 73.2% of the cohort met criteria for a history of at least one assessed psychiatric disorder, the most common of which were MDD, ADHD, oppositional defiant disorder (ODD), conduct disorder, and post-traumatic stress disorder (PTSD). Interestingly, a small number of youth were found to already meet criteria for a bipolar (*n* = 20) or psychotic spectrum disorder (*n* = 7), all of whom had been enrolled as a high SMI-risk participant. Despite the relatively young age of the cohort, a number of youth also met criteria for a tobacco (*n* = 69), alcohol (*n* = 53), or other substance use disorder (*n* = 77), the majority of whom were enrolled as a high SMI-risk participant.

**Table 3.** Rates of lifetime history of psychiatric disorders assessed via the Kiddie Schedule for Affective Disorders and Schizophrenia – Computerized Version (KSADS-COMP) for overall currently enrolled cohort and comparison of high versus lower serious mental illness (SMI) risk groups.

| KSADS-COMP Diagnosis | Total Sample<br>$n$ (%) | High SMI-Risk<br>$n$ (%) | Low SMI-Risk<br>$n$ (%) | Odds Ratio [95% CI] | p-value | FDR<br>corrected<br>q-value |
| --- | --- | --- | --- | --- | --- | --- |
| Umbrella Depressive Disorders | 667 (28%) | 642 (29%) | 25 (14.5%) | 2.26 [1.42, 3.59] | 0.001 | 0.004 |
| Major Depressive Disorder | 666 (27.9%) | 641 (29%) | 25 (14.5%) | 2.25 [1.41, 3.58] | 0.001 | 0.004 |
| Persistent Depressive Disorder | 93 (3.9%) | 88 (4%) | 5 (2.9%) | 1.12 [0.44, 2.85] | 0.813 | 0.996 |
| Disruptive Mood Dysregulation Disorder | 26 (1.1%) | 25 (1.1%) | 1 (0.6%) | 1.86 [0.25, 13.86] | 0.546 | 0.901 |
| Umbrella Bipolar Disorders | 20 (0.8%) | 20 (0.9%) | 0 (0%) | 2606792.81 [0.00, Inf] | 0.985 | 0.996 |
| Bipolar Type 1 | 18 (0.8%) | 18 (0.8%) | 0 (0%) | 6033584.83 [0.00, Inf] | 0.99 | 0.996 |
| Bipolar Type 2 | 2 (0.1%) | 2 (0.1%) | 0 (0%) | 2394145.92 [0.00, Inf] | 0.995 | 0.996 |
| Umbrella Psychotic Disorders | 7 (0.3%) | 7 (0.3%) | 0 (0%) | 1724665.72 [0.00, Inf] | 0.991 | 0.996 |
| Schizophrenia | 3 (0.1%) | 3 (0.1%) | 0 (0.0%) | 3148389.19 [0.00, Inf] | 0.996 | 0.996 |
| Schizophreniform Disorder | 0 (0.0%) | 0 (0.0%) | 0 (0.0%) | -- | -- | -- |
| Schizoaffective Disorder | 4 (0.0%) | 4 (0.2%) | 0 (0.0%) | 3422454.82 [0.00, Inf] | 0.994 | 0.996 |
| Umbrella Anxiety Disorders | 525 (22%) | 507 (22.9%) | 18 (10.4%) | 2.42 [1.47, 4.00] | 0.001 | 0.004 |
| Panic Disorder | 50 (2.1%) | 49 (2.2%) | 1 (0.6%) | 3.55 [0.49, 25.99] | 0.212 | 0.496 |
| Agoraphobia | 27 (1.1%) | 26 (1.2%) | 1 (0.6%) | 1.85 [0.25, 13.81] | 0.547 | 0.901 |
| Separation Anxiety | 211 (8.9%) | 199 (9%) | 12 (6.9%) | 1.36 [0.74, 2.50] | 0.321 | 0.600 |
| Social Anxiety Disorder | 162 (6.8%) | 155 (7%) | 7 (4%) | 1.58 [0.72, 3.44] | 0.254 | 0.507 |
| Generalized Anxiety Disorder | 237 (9.9%) | 235 (10.6%) | 2 (1.2%) | 9.37 [2.30, 38.21] | 0.002 | 0.010 |
| Obsessive-Compulsive Disorder | 43 (1.8%) | 43 (1.9%) | 0 (0%) | 5938697.13 [0.00, Inf] | 0.985 | 0.996 |
| Attention-Deficit/Hyperactivity Disorder | 899 (37.7%) | 869 (39.3%) | 30 (17.3%) | 3.07 [2.03, 4.63] | 1.07E-07 | 2.991e-06 |
| Oppositional Defiant Disorder | 523 (21.9%) | 503 (22.7%) | 20 (11.6%) | 2.04 [1.26, 3.30] | 0.004 | 0.017 |
| Conduct Disorder | 324 (13.6%) | 309 (14%) | 15 (8.7%) | 1.40 [0.81, 2.44] | 0.23 | 0.496 |
| Autism Spectrum Disorder | 63 (2.6%) | 62 (2.8%) | 1 (0.6%) | 4.88 [0.67, 35.49] | 0.117 | 0.365 |
| Umbrella Trauma Stress Disorder | 335 (14.1%) | 319 (14.4%) | 16 (9.2%) | 1.56 [0.91, 2.68] | 0.103 | 0.361 |
| Post-Traumatic Stress Disorder | 313 (13.1%) | 297 (13.4%) | 16 (9.2%) | 1.41 [0.82, 2.42] | 0.213 | 0.496 |
| Adjustment Disorder | 27 (1.1%) | 27 (1.2%) | 0 (0%) | 4368483.26 [0.00, Inf] | 0.985 | 0.996 |
| Umbrella Substance Use Disorder | 127 (5.3%) | 126 (5.7%) | 1 (0.6%) | 7.50 [1.02, 54.97] | 0.047 | 0.190 |
| Tobacco Use Disorder | 69 (2.9%) | 68 (3.1%) | 1 (0.6%) | 3.77 [0.51, 27.77] | 0.193 | 0.496 |
| Alcohol Use Disorder | 53 (2.2%) | 53 (2.4%) | 0 (0%) | 4061284.76 [0.00, Inf] | 0.983 | 0.996 |
| Other Substance Use Disorders | 77 (3.2%) | 74 (3.3%) | 3 (1.7%) | 1.25 [0.38, 4.17] | 0.713 | 0.996 |
| Participants without a Diagnosis | 640 (26.8%) | 555 (25.1%) | 85 (49.1%) | -- | -- | -- |

Between youth recruited for high versus lower-SMI risk, logistic regression models showed significantly elevated rates of any depressive disorder (odds ratio (OR) = 2.26, 95%CI[1.42, 3.59], *p* = .001, *q* = .004), MDD (OR = 2.25, 95%CI[1.41, 3.58], *p* = .001, *q* = .004), any anxiety disorder (OR = 2.42, 95%CI[1.47, 4.00], *p* = .001, *q* = .004), GAD (OR = 9.37, 95%CI[2.30, 38.21], *p* = .002, *q* = .010); ADHD (OR = 3.07, 95%CI[2.03, 4.63], *p* < .001, *q* < .001), and ODD (OR = 2.04, 95%CI[1.26, 3.30], *p* = .004, *q* = .017) among high SMI-risk compared to lower SMI-risk youth (Table 3).

#### Adverse Childhood Experiences

Table 4 shows the rates of ACEs among currently enrolled participants, and for youth recruited for high-versus lower-SMI risk. The most common ACEs across youth were: having parents who were separated (57.8%), having lived with a family member with mental illness (49.6%) or who used substances (36.2%), witnessing domestic violence (34.9%), and experiencing bullying (34.5%).

**Table 4.**
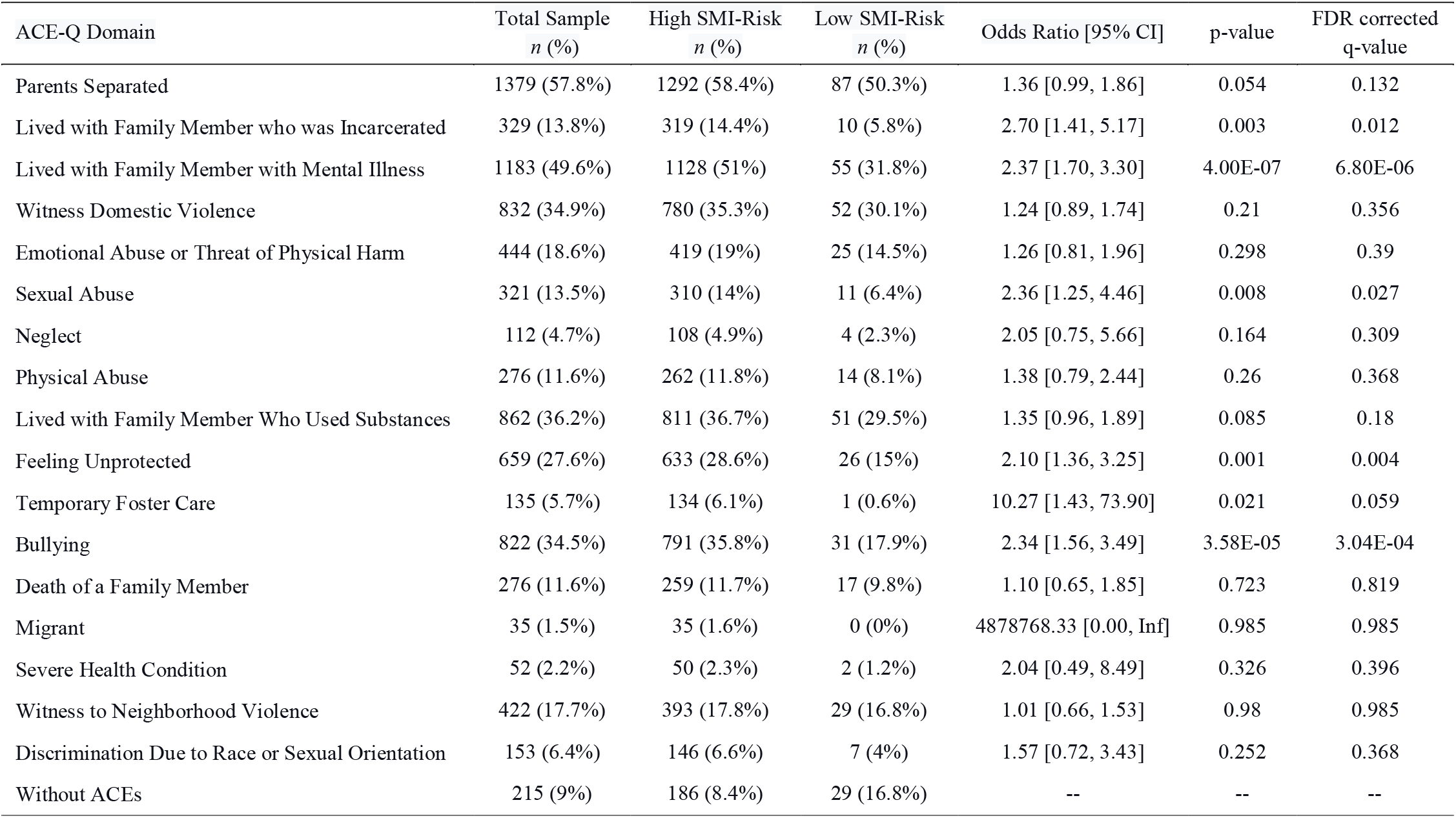
Rates of adverse childhood experiences as assessed by the Adverse Childhood Experiences Questionnaire (ACE-Q) for overall currently enrolled cohort and comparison of high versus lower serious mental illness (SMI) risk groups.

| ACE-Q Domain | Total Sample<br>n (%) | High SMI-Risk<br>n (%) | Low SMI-Risk<br>n (%) | Odds Ratio [95% CI] | p-value | FDR corrected<br>q-value |
| --- | --- | --- | --- | --- | --- | --- |
| Parents Separated | 1379 (57.8%) | 1292 (58.4%) | 87 (50.3%) | 1.36 [0.99, 1.86] | 0.054 | 0.132 |
| Lived with Family Member who was Incarcerated | 329 (13.8%) | 319 (14.4%) | 10 (5.8%) | 2.70 [1.41, 5.17] | 0.003 | 0.012 |
| Lived with Family Member with Mental Illness | 1183 (49.6%) | 1128 (51%) | 55 (31.8%) | 2.37 [1.70, 3.30] | 4.00E-07 | 6.80E-06 |
| Witness Domestic Violence | 832 (34.9%) | 780 (35.3%) | 52 (30.1%) | 1.24 [0.89, 1.74] | 0.21 | 0.356 |
| Emotional Abuse or Threat of Physical Harm | 444 (18.6%) | 419 (19%) | 25 (14.5%) | 1.26 [0.81, 1.96] | 0.298 | 0.39 |
| Sexual Abuse | 321 (13.5%) | 310 (14%) | 11 (6.4%) | 2.36 [1.25, 4.46] | 0.008 | 0.027 |
| Neglect | 112 (4.7%) | 108 (4.9%) | 4 (2.3%) | 2.05 [0.75, 5.66] | 0.164 | 0.309 |
| Physical Abuse | 276 (11.6%) | 262 (11.8%) | 14 (8.1%) | 1.38 [0.79, 2.44] | 0.26 | 0.368 |
| Lived with Family Member Who Used Substances | 862 (36.2%) | 811 (36.7%) | 51 (29.5%) | 1.35 [0.96, 1.89] | 0.085 | 0.18 |
| Feeling Unprotected | 659 (27.6%) | 633 (28.6%) | 26 (15%) | 2.10 [1.36, 3.25] | 0.001 | 0.004 |
| Temporary Foster Care | 135 (5.7%) | 134 (6.1%) | 1 (0.6%) | 10.27 [1.43, 73.90] | 0.021 | 0.059 |
| Bullying | 822 (34.5%) | 791 (35.8%) | 31 (17.9%) | 2.34 [1.56, 3.49] | 3.58E-05 | 3.04E-04 |
| Death of a Family Member | 276 (11.6%) | 259 (11.7%) | 17 (9.8%) | 1.10 [0.65, 1.85] | 0.723 | 0.819 |
| Migrant | 35 (1.5%) | 35 (1.6%) | 0 (0%) | 4878768.33 [0.00, Inf] | 0.985 | 0.985 |
| Severe Health Condition | 52 (2.2%) | 50 (2.3%) | 2 (1.2%) | 2.04 [0.49, 8.49] | 0.326 | 0.396 |
| Witness to Neighborhood Violence | 422 (17.7%) | 393 (17.8%) | 29 (16.8%) | 1.01 [0.66, 1.53] | 0.98 | 0.985 |
| Discrimination Due to Race or Sexual Orientation | 153 (6.4%) | 146 (6.6%) | 7 (4%) | 1.57 [0.72, 3.43] | 0.252 | 0.368 |
| Without ACEs | 215 (9%) | 186 (8.4%) | 29 (16.8%) | -- | -- | -- |

Youth recruited for high SMI-risk had significantly higher rates of having lived with a family member who was incarcerated (OR = 2.70, 95%CI[1.41, 5.17], *p* = .003, *q* = .012) or who had a mental illness (OR = 2.37, 95%CI[1.71, 3.30], *p* < .001, *q* < .001), having experienced sexual abuse (OR = 2.36, 95%CI[1.25, 4.46], *p* = .008, *q* = .027), feeling unprotected (OR = 2.10, 95%CI[1.36, 3.25], *p* = .001, *q* = .004), and experiencing bullying (OR = 2.34, 95%CI[1.56, 3.49], *p* < .001, *q* < .001), compared to youth at lower SMI-risk. Few participants reported no ACEs (9.0%).

## Discussion

With a target sample of 3,000 children and adolescents, this study will establish one of the largest cohorts of youth at transdiagnostic high-risk for SMI involving high-quality deep phenotyping, genome-wide genotyping, and longitudinal follow-up. Indeed, passive longitudinal monitoring of the cohort via EMR is one of the most promising aspects of the study, as this will allow long-term mental health trajectories to be linked with data on early clinical and neurobehavioral functioning and environmental exposures, in addition to genetic risk. Furthermore, having both family- and EMR-based high-risk recruitment streams will facilitate future analyses comparing the utility of different indices of risk (e.g., family-, clinical-, environmental-vs. molecular genetic-based risk) for predicting later SMI outcomes, a critical area of interest.

This cohort aims to be the largest SMI-risk youth cohort of this size in Latin America, contributing critically to improved representation of non-European ancestry individuals in genetic and psychiatric research. It also represents a unique opportunity to study developmental trajectories in a population in a LMIC. While Latino individuals are represented to a degree in existing youth cohorts in North America and Europe, such as in the ABCD Study, which is a large, longitudinal, general population study of brain development in the United States that includes genetic, behavioral, and clinical information, greater exposure to major environmental stressors in Colombia allows broader questions related to adversity and resilience to be investigated.^8^ For example, while in Latin migratory contexts in North America, stressful experiences related to discrimination^43^, cultural displacement^44,45^, and job insecurity are common^46^, in Antioquia, elevated rates of forced displacement, armed violence, structural poverty, and substance use are added.^5,47^ Parental separation, domestic violence, and substance use in the family environment were some of the most frequently reported adversities among currently enrolled youth, along with bullying. Although physical and sexual abuse were reported in a somewhat lower portion of youth, these are strong risk factors for psychopathology.^48,49^ However, the fact that some children exposed to these experiences have not yet presented with mental disorders suggests the presence of resilience mechanisms that are important to investigate further. Consistent with our ascertainment strategies, there are elevated rates of ACEs in this cohort compared to the general population ABCD Study in the U.S. For example, in the ABCD Study cohort, at year 2, when youth were 11-12 years of age and therefore similar to the average age in the current cohort, 17.20% of the cohort had experienced no ACEs^50^ compared to 9.0% of the current cohort. Similarly, rates of many specific ACEs such as physical abuse (1.07%), sexual abuse (1.22%), and household divorce or separation (17.24%) were lower in the ABCD Study cohort at year 2^50^, than in the current cohort (i.e., rates of 11.6%, 13.5%, and 57.8%, respectively, for the same ACEs). Longitudinal study of this cohort offers important opportunities to understand how adversity and protective factors contribute to the development of, or resilience to, psychopathology and SMI, not only in Colombia, but world-wide.

Interestingly, the rates of MDD, ADHD, ODD, CD, and PTSD, and other diagnoses among high SMI-risk youth in the current study are higher compared to a large study in Brazil which also used dual recruitment streams of elevated family psychopathology and psychopathology in children to establish a high-risk youth cohort.^51^ This study found a prevalence of any current mental disorder in high-risk children of 29.7%. In contrast, the rate of a history of any mental disorder in the familial high-risk SMI group in the current “Paisa Kids” study was 39.5%. Although rate differences likely reflect, at least in part, the difference between current versus lifetime psychiatric history rates, elevated psychopathology in the current cohort is likely to facilitate well-powered investigations into developmental factors associated with distinct SMI outcomes over time.

Taken together, our large sample of youth enriched transdiagnostically for SMI risk with dual family-based and EMR-based recruitment streams; rich clinical, neurobehavioral, and environmental data; elevated ACE rates; and passive monitoring of longitudinal clinical outcomes, offers critical opportunities to identify differentiated trajectories for distinct SMI outcomes and improve early detection of SMI risk. Additional questions will be able to be investigated as data are shared with the scientific community through the NIH Data Archive. Ultimately, this project offers promise to advance us towards a more preventive, equitable, and contextualized child psychiatry, capable of better responding to mental health challenges in more diverse vulnerable populations.

## Data Availability

All data produced in the present study will be available to authors with approved access through the NIH Data Archive.

https://dx.doi.org/10.15154/v3a4-mt40

