## Supplementary Information for "The Paisa Kids Study: Rationale, Methods, and Initial Characterization of a Youth Cohort at Transdiagnostic High Risk for Serious Mental Illness"

***Mental Health, Behavioral Traits, and Psychosocial Functioning***

**Kiddie Schedule for Affective Disorders and Schizophrenia - Computerized (KSADS-COMP).** The Spanish version of the computer-assisted Kiddie Schedule for Affective Disorders and Schizophrenia for School-Aged Children (KSADS-COMP) structured clinical interview was used to obtain psychiatric diagnosis history at study entry. The KSADS is the most widely used and well-validated diagnostic interview for children and adolescents.^1^ The KSADS-COMP automatically scores and branches the interview and classifies responses to identify current and past history of each DSM-5 psychiatric disorder, streamlining administration and data entry.^1,2^

Primary caregivers and the minor were interviewed separately. Caregivers completed the following modules about the minor: mood disorder, psychosis, panic disorder, agoraphobia, separation anxiety, social anxiety disorder, GAD, obsessive compulsive disorder, ADHD, oppositional defiant disorder, conduct disorder, ASD, alcohol use disorder, drug use disorder, post-traumatic stress disorder, sleep problems, and suicidality. Children and adolescents completed the modules for: mood disorder, psychosis, conduct disorder, alcohol use disorder, drug use disorders, and post-traumatic stress disorder. The minor’s history of mental health and alcohol and/or drug addiction service use, including psychotherapy and medication, was also assessed by the KSADS-COMP.

**Child Behavior Checklist (CBCL) Children's Dimensional Scale.** The CBCL is a widely used and well-validated 138-item measure of behavioral and emotional functioning in children and adolescents aged 6 to 18.^3,4^ In addition to a total score and scores for internalizing and externaling problems, the CBCL yields scores for several syndromic scales, including anxiety/depression, somatic complaints, social problems, thinking problems, attention problems, rule-breaking behavior, and aggressive behavior. Respondents respond on a 3-point likert scale to whether each statement presented is “Not True”, “Somewhat or Sometimes True”, or “Very True or Often True”. The 113 questions used to assess psychopathology and behavioral problems were utilized in the current study. The 25 optional questions used to derive competence scales for activities, social, and school functioning were not administered, as these areas were partially assessed during the KSADS. Caregivers completed the CBCL for youth of all ages. Adolescents, aged 12 and up additionally completed the self-report version of the scale. Example items include:

1) Acts too young for age (Actúa como si fuera mucho menor que su edad)

2) Drinks alcohol without parents approval (Toma bebidas alcohólicas sin permiso de los padres).

**Anhedonia scale.** The anhedonia scale is a 14-item questionnaire for adolescents that quantifies anhedonia severity in the past two weeks in adolescents,^5^ including social versus physical anhedonia. The scale has acceptable to excellent internal consistency (α = .79 - .94) and modest test-retest reliability (ICC = .73).^5^ Participants indicate on a 4-point likert scale to what extent each statement applies to them, with responses ranging from Never (1) to Always (4). Example items include:

1) I had no motivation to get started on things (No tuve motivación para empezar a hacer las cosas.)

2) Nothing made me feel excited (Nada me hizo sentir emocionado.)

**Big Five Questionnaire for Children (BFC-SF).** The 20-item Big Five Questionnaire for Children - Short Form (BFC-SF)^6^ was used to assess the five dimensions of the Big Five model of personality: openness to experience, conscientiousness, extraversion, agreeableness, and neuroticism. Children report their agreement with statements related to these dimensions on a 0-2 likert scale (0 = not true to 2 = certainly true). It has shown acceptable to good internal consistency across studies and subscales (α = .64 - .86).^7,8^ Example items include:

1) I like to be with other people (Me gusta estar con otras personas)

2) I get upset easily (Me molesto fácilmente)

***Neurobehavioral Functioning***

**NIH Toolbox for Neurological and Behavioral Functioning (NIH-TB).** The NIH-TB is an expert-curated compilation of assessment measures spanning cognitive, sensory, motor, and emotional functioning. It was developed as part of the NIH Blueprint for Neuroscience Research to create a streamlined, “common currency” for assessing key neurobehavioral constructs across developmental stages. Most measures can therefore be administered in children as young as 3 years of age, and norms exist for individuals aged 3-85. A Spanish version is available and has been validated^9,10^ and administration occurs via an iPad app to facilitate ease of administration. The cognitive battery and select tests from the sensory and motor battery were administered to all participants.

***Cognitive Function: NIH-TB Cognitive Battery.*** All subtests from the main NIH-TB cognitive battery were administered, allowing scores for individual subtests and a total composite score to be generated. Specifically, the Dimensional Change Card Sort Test, which measures the set-shifting component of executive function, Flanker Inhibitory Control and Attention Test, which measures the attentional component of executive function, Picture Sequence Memory Test which measures episodic memory, Picture Vocabulary and Oral Reading Recognition Tests, which each measure language, Pattern Comparison Test, which measures processing speed, and Word List Sorting Test, which measures working memory, were administered.

Two-week test-retest reliability for the cognitive total composite score was strong among youth (ICC values ≥ .88)^11^, and internal consistency estimates for cognitive summary scores were also high.^12^

***Sensory Function: NIH-TB Words-in-Noise Test.*** The Words-in-Noise Test, which assesses one’s ability to perceive words in a noisy environment, was used to assess sensory function. Participants are asked to repeat out loud the word they hear in each trial of a recording, where a single word is presented in the presence of increasingly louder background noise. Moderate test-retest reliability in Spanish populations has been demonstrated (Spearman's test coefficients = .49 and .55 for the left and right ear, respectively).^9^

***Motor Function: NIH-T 9-Hole Pegboard Dexterity Test - C.*** The 9-Hole Pegboard Test is a commonly used and well-validated timed assessment of fine motor ability^13^, in which participants sequentially place cylindrical pegs into a 3 by 3 matrix of holes, followed by removing them. Two trials are administered per dominant and non-dominant hand.

**Rey Auditory Verbal Learning Test.** The Rey Auditory Verbal Learning Test is a widely used assessment of short-term verbal learning and memory that has shown good test-retest reliability and internal consistency (ICC of .76 and α = .84, respectively).^14^ The Spanish version used for the present study was generated by the CENTER-TBI Consortium using linguistically and culturally adapted translations of original English source lists, with the aim of retaining characteristics such as the familiarity, concreteness and imageability of words.^15^ Three immediate memory trials were administered for the current study.

***Environment***

**Adverse Childhood Experiences (ACEs) Questionnaire.** ACEs were assessed using the 17-item ACE questionnaire, which covers multiple domains including physical, emotional, and sexual abuse, physical and emotional neglect, substance abuse or mental illness in a family member, parental separation or divorce, and domestic violence^16^. It has good internal consistency (α = .70).^17^ Example questions include:

1) Your child’s parents or guardians were separated or divorced (Los padres (o tutores) de su hijo están separados o divorciados).

2) Your child lived with a household member who served time in jail or prison (Su hijo vivió con un miembro del hogar que está o estuvo condenado en la cárcel o prisión).

**Questionnaire of Unpredictability in Childhood - Adolescents/Parents.** Unpredictability in early childhood was measured with the Questionnaire of Unpredictability in Childhood - Spanish - Brief (QUIC-SP-5), which is a 5-item measure derived from the original 38-item QUIC, that showed strong correlations with the original full version (*r’s* = 0.81–0.87) and predicted internalizing symptoms with similar effect sizes as the full version.^18^ The QUIC-SP-5 scores range from 0 to 5, with higher scores indicating greater unpredictability. Respondents indicate whether each statement is true/false. Example items include:

1) Before age 11, my child has/had a bedtime routine (e.g., was tucked in, parents read him/her a book, took a bath). (Antes de los 11 años, mi hijo tiene / tenía una rutina a la hora de acostarse (por ejemplo, era acobijado, los padres le leyeron un libro, se bañaba)

2) At least one of my child’s parents is unpredictable. (Al menos uno de los padres de mi hijo es impredecible).

**Child Promotive Factors - Adolescents/Parents.** Questions on promotive factors for youth health across individual-, family-, and neighborhood-levels were adapted from the United States National Survey of Children’s Health Questionnaires for ACEs and Promotive Factors, similar to others ^19^. Specific included questions were:

1) During the past week, on how many days did all the family members who live in the household eat a meal together?

a) 0 days
b) 1-3 days
c) 4-6 days
d) Every Day

2) During the past 12 months, did the child participate in a sports team or classes after school or on weekends; a club or organization after school or on weekends; other organized activities or lessons such as music, dance, language or other arts?

a) Yes
b) No

3) During the past 12 months, did the child participate in any type of community service or volunteer work at school, place of worship, or in the community?

a) Yes
b) No

4) To what extent to you agree with these statements about your neighborhood or community: People in this neighborhood help each other out

a) Definitely Agree
b) Somewhat Agree
c) Somewhat Disagree
d) Definitely Disagree

5) In general, how well can you and this child share ideas or talk about things that really matter

a) Definitely Agree
b) Somewhat Agree
c) Somewhat Disagree
d) Definitely Disagree

1) DURANTE LA SEMANA PASADA, ¿cuántos días comieron juntos una comida, todos los miembros de la familia que conviven en el hogar?

a) 0 días
b) 1-3 días
c) 4-6 días
d) Todos los días

2) DURANTE LOS ÚLTIMOS 12 MESES, ¿participó este niño en un equipo deportivo o tomó clases deportivas después de la escuela o en fines de semana? ¿Un club u organización después de la escuela o los fines de semana? ¿Otras actividades o lecciones organizadas, como música, danza, lenguaje u otras artes?

a) Sí
b) No

3) DURANTE LOS ÚLTIMOS 12 MESES, ¿participó este niño en cualquier tipo de servicio comunitario o trabajo voluntario en la escuela, lugar de culto, o en la comunidad?

c) Sí
d) No

4) ¿Hasta qué punto está usted de acuerdo con las siguientes afirmaciones sobre su vecindario o comunidad?

a) De acuerdo
b) Algo de Acuerdo
c) Algo en Desacuerdo
d) Definitivamente en Desacuerdo

***Obstetric, Developmental, and Medical History***

**Lewis-Murray Obstetric Complications Scale - Parent.** Obstetric history was assessed using the Lewis-Murray scale, which is a commonly used, 15-item retrospective recall measure organized into three subscales: 1) pregnancy complications (i.e., maternal infection, preeclampsia, antepartum hemorrhaging or threatened misscarriage during pregnancy); 2) abnormal fetal growth (i.e., low birthweight, complications delivering twins, severe physical abnormalities with child); and 3) development and delivery difficulties (i.e., pre-rupturing of membranes, prolonged labor, delivery complications caused by umbilical cord, presence of cesarean section, complication, or emergency, abnormal presentation, newborn was in incubator.^20,21^ Scores are on a 3-point likert scale for absent (0), equivocal (1), or definite (2). Comparison of maternal recall for complications rated as “definite” to medical records have shown strong agreement for most items (Overall kappa = .65).^22,23^
